# Predicting Success of Phase III Trials in Oncology

**DOI:** 10.1101/2020.12.15.20248240

**Authors:** Stephan Hegge, Markus Thunecke, Matthias Krings, Léonard Ruedin, Jan Saputra Müller, Paul von Bünau

**Affiliations:** Hegge Holding UG, Kopernikusstr. 24, 10247 Berlin; Catenion GmbH, Münzstrasse 18, 10278 Berlin, Germany; idalab GmbH, Potsdamer Straße 68, 10785 Berlin, Germany; AskBy GmbH, Boxhagener Str. 18, 10245 Berlin

## Abstract

**Importance:** We developed a model predicting the probability of success (PoS) for single planned or ongoing PhIII trials based on information available at trial initiation. Such a model is highly relevant for study sponsors to capture risk and opportunity on a trial-to-trial basis through trial optimization, and for investors to select drugs whose trial design match their investment strategy.

**Objectives:** To predict the outcome of planned or ongoing PhIII trials in oncology, given publicly available prior information

**Design, Setting, Participants:** Predictive modeling using publicly available data for 360 completed PhIII and 1240 PhII studies initiated between 2003 and 2012. Success and failure of PhIII studies were modeled using Bayesian logistic regression model.

**Main Outcome Measures:** Predicted PoS of individual PhIII trials based on a Bayesian model calibrated on publicly available data translated into 16 composite scores. Those scores cover aspects such as trial design, indication, number of patients, phase II (PhII) study outcomes, experience of sponsor at time of trial initiation, and others.

**Results:** The model allows to calculate the PoS distribution – including credible intervals – for a PhIII trial in oncology. The predictive performance was determined using an area under the receiver-operator curve (AUROC), resulting in an overall performance of 73%_oPP_ (mean AUROC). We identified two key factors contributing to the predictive performance of the model: quality and strength of PhII data and experience of the sponsor at the time of study initiation.

**Conclusion and Relevance:** We describe the generation and application of a statistical model predicting the PoS for individual PhIII trials in oncological indications with unprecedented predictive performance. Compared to other approaches, this is the first study generating a fully transparent model resulting in trial-specific PoS distributions. Moreover, we have shown that qualitative concepts such as PhII knowledge or sponsor R&D strength can be captured in quantitative scores and that these scores have a high predictive power.

**Key Points:** *Question:* What is the probability of success (PoS) for single phase III (PhII)I trials in oncology?

*Findings:* We developed a model allowing to predict the PoS of single PhIII trials in oncology with a predictive performance of 73%_PP_ and demonstrated that qualitative factors such as strength of PhII knowledge and sponsor R&D strength can be captured in quantitative scores that have significant predictive power.

*Meaning:* The model can help study sponsors to analyze and amend planned clinical trials, and investors to choose where to invest best.

## Introduction

In recent years, predictive algorithms have become ubiquitous across a wide range of industries, such as logistics – e.g. Amazon’s predictive shipping^1^ – or information retrieval – e.g. Google’s predictive search algorithm^2^. By combining information from a vast number of sources in an objective, unbiased manner, predictive algorithms can outperform human decision making with respect to accuracy and speed at marginal cost. Even in the public sector, political decision makers have become increasingly aware of the importance of accurate predictions and have started evaluating different approaches in forecasting tournaments^3^. Predictive algorithms can aid decision makers in the pharmaceutical industry as well. However, so far, adoption has been limited. The current decision making process, from discovery to clinical development phases, is characterized by a series of decision points defined by formal go/no-go criteria^4^ that relate to the available clinical data. This process is implemented to aid executives as it reflects regulatory requirements for each indication. However, it has been demonstrated that decisions based on real world cases vary greatly, ranging from absolutely ‘go’ to absolutely ‘no-go’ due to subjective interpretation of identical data^5^.

We argue that predictive algorithms can be employed for improving and rationalizing decision making in Pharma, particularly in clinical trials representing the most crucial and expensive centerpiece of drug development. Predictive algorithms based on Big Data have demonstrated that they are able to aid or even outperform drug developers and physicians when it comes to predicting either patient accrual rates in clinical trials^6,7^, or optimal cancer rehabilitation^8^, or supportive care interventions^8^.

Trial decision making for any drug in clinical development has far-reaching implications. On one hand, hundreds to thousands of patients are recruited to test the effects, each of them hoping to benefit from the new drug. On the other hand, sponsors risk tens to hundreds of million Dollars on trials that may or may not demonstrate the drug’s effect^9^. It is therefore in the interest of sponsors to terminate failures early, without compromising on the quality of clinical trials or terminating successful agents, and – most importantly – without exposing clinical trial subjects to unnecessary risk^10^. It is also in the interest of patients to participate in trials that have a strong chance to achieve a positive outcome.

Success and failure rates in pharmaceutical drug development are described by several sources, ranging from commercial benchmarking providers like BioMedTracker ^11,12^ to academic papers ^13–16^.

Still, no established method exists to predict the probability of success (PoS) for an individual clinical trial; in fact, there is no clear definition for trial success and failure. Hence current approaches^11–15^ focus on determining the average attrition rates from phase to phase – so called phase transitions counts (PTC) – during drug development resulting in historical success rates (hSRs). Phase transitions – and hence hSRs – are either assessed on an indication^11^ (hSR_Indication_; eFig 1, eTable 1), or on a program level^10,17^ (hSR_Program_, eFig 1, eTable 1). It has become gold standard in trial planning and portfolio management to take hSRs and use them as forward-looking estimates of PoS. Moreover, using hSR as gauge for forward-looking PoS is limited to factors that allow for straightforward stratification. Drivers that are more complex in nature, such as the knowledge a sponsor has accumulated from prior phases, cannot be analysed using simple hSRs. This approach has far-reaching implications in decision making for several reasons:

**Figure 1.**
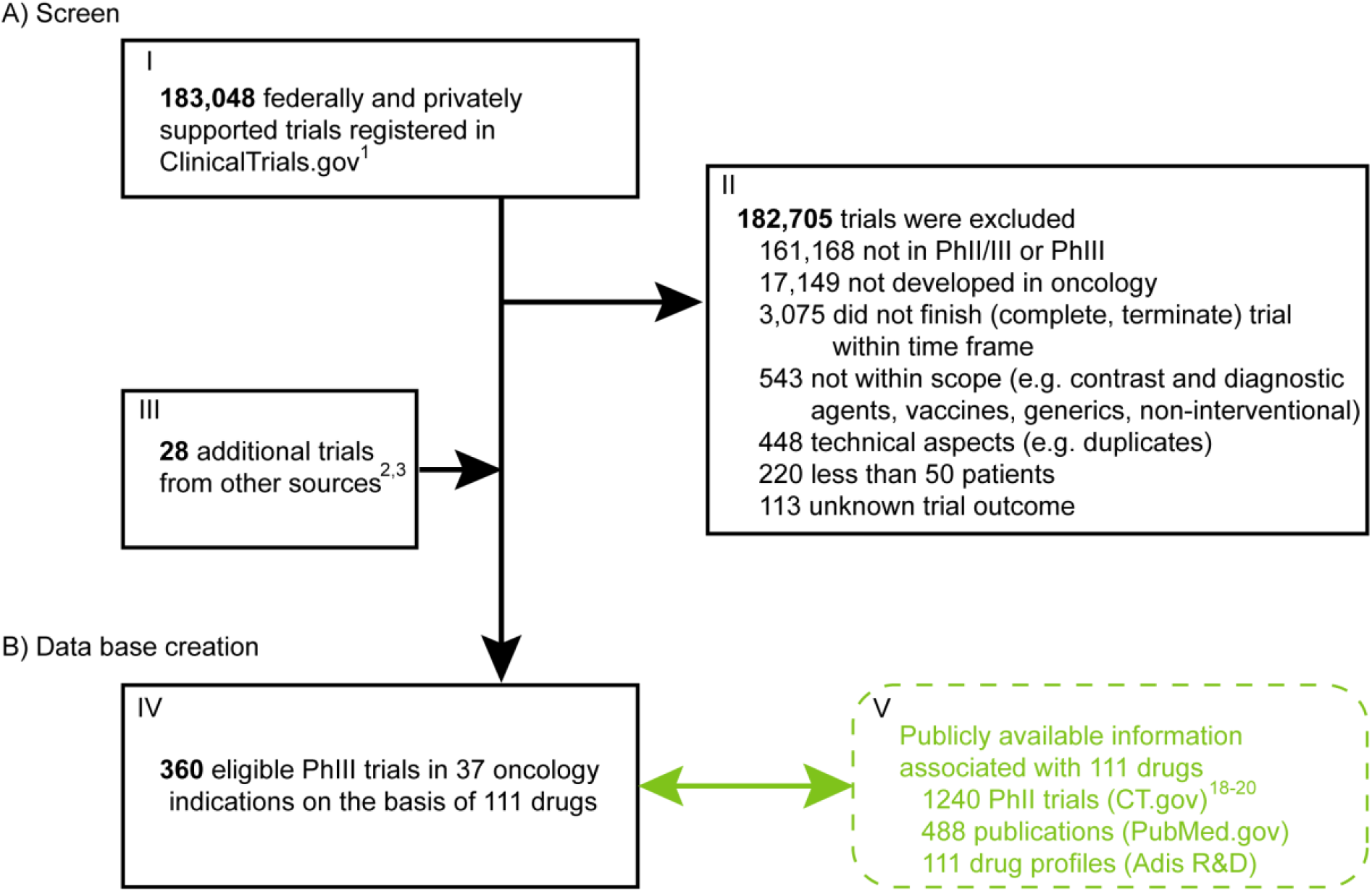
Consort flow diagram. (A) Screen. 360 eligible PhIII studies were identified screening ClinicalTrials.gov^18^ and other clinical trial registries^19,20^ (I-IV, black font). (B) Database creation. Associated information for each eligible PhIII trial was searched for and added using public sources (V, green font) to create a project specific database^18–22^ (IV).

a. hSRs are historic, past transition rates, however they are naively used as forward-looking probabilities, without a statistical evaluation of their predictive power
b. hSRs are averages. Thus, applying these as forward-looking PoS to specific individual trials will systematically overestimate the PoS of riskier than average drug development programs, while underappreciating the PoS of particularly well conducted low-risk programs
c. hSR-based PoS values are typically used as point estimates without credible intervals^11^. This can lead to a false sense of confidence, and falsely informed decisions – such as terminating a drug that would work and be safe in favor of continuing a drug that is ineffective or unsafe – when rating the PoS of one program higher than another
d. Neither of the hSR approaches consider individual trials, but are limited to indications^11^, drug programs^10^ or even entire therapeutic areas^17^

Here we present an approach providing forward looking PoS estimates for individual PhIII trials in oncology based on publicly available data. Firstly, all potentially relevant parameters were classified and translated into numerical or categorical data. Secondly, we identified and quantified correlations between each parameter and trial outcome. Thirdly, we developed and calibrated a Bayesian algorithm for predicting trial outcomes, which provides a full posterior distribution. Fourthly, we have shown that complex qualitative factors such as PhII knowledge or sponsor R&D strength can be modeled using quantitative scores and that these scores are indeed significantly predictive. This approach allows decision-makers to capture risk and opportunity on a trial-to-trial basis, and helps trial sponsors and planners to identify and mitigate trial- and indication-specific risks.

## Materials and Methods

### Data Sources

ClinicalTrials.gov (CT.gov) is a publicly accessible database run by the United States National Library of Medicine at the National Institutes of Health^18^. It served as a starting point for our analysis as it is the largest registry for clinical trials to date providing general characteristics and outcome information of about ∼283,000 (as of January 2018) federally and privately supported clinical trials. 47% of studies recruit patients outside the US, 36% of trials are recruiting in the US only, and 6% recruit patients in both, US and non-US locations, while 11% do not provide that information ^18^.

To complement for potentially publicly available information not listed on CT.gov we used the European Clinical Trial Database^19^, JAPIC Clinical Trials Information^20^, PubMed^21^ and Adis R&D^22^ (ADIS). We used PubMed to identify publications associated with each given trial. ADIS is a database for drug development gathering all available information (*in vitro* and *in vivo* experiments, preclinical data, clinical trial outcomes, chemistry, conference posters, conference abstracts, press releases etc.) for a given compound.

### Study Sample

We queried the database of CT.gov^18^ (>183,000 trials by April 2014) and applied filters to identify a sample of novel therapeutics (i.e., new chemical entities (NCEs) or novel biological entities (NBEs)) for which a PhIII study in oncology had been initiated between 01.01.2003 and 01.03.2012 (Fig 1). Filter criteria excluded generics, biosimilars, reformulations, radiotherapy, and combination therapies of two or more non-novel therapeutics. Cell-therapies, non-therapeutic agents, such as diagnostic tools, contrast agents and supportive care approaches were also not considered. We removed all duplicated records.

### Technical Implementation

For MCMC sampling, we used the software package JAGS version 4.3. All other computations were performed in Python version 2.7, relying heavily on pandas version 0.17.

## Results

### Database

ClinicalTrials.gov is the largest registry of clinical trials database with >183,000 registered trials at the time of query (Fig 1, I). Strict application of exclusion criteria resulted in 360 PhIII trials across 37 oncology indications (Fig 1, IV). The 111 NBEs and NCEs associated with these PhIII studies served as a starting point for the complementary analysis, which aimed to gather all publicly available PhII and PhIII information associated with the original set of 360 PhIII studies by means of searching for the drug name, the trial identifier, trial names and its synonyms. We searched in the European and Japanese trial registries EudraCT and JAPIC, scanned published literature using PubMed.gov, company homepages, and checked Adis R&D, a database for drug research and development (Fig 1, V). This approach gave rise to 1240 PhII studies, 488 publications, and 111 ADIS data entries, respectively, with detailed information about PhII and PhIII study outcomes, endpoint measures, study design, actual patient population etc. (eTables 1-3). Please note, PhI results were not considered for this proof-of-principle analysis, given PhI studies are less well defined with regard to patient stratification (mixed patient population with regard to tumor type, tumor stage, line of treatment) and endpoint measures. We defined trial success by meeting all primary endpoints (see eTable 3 for detailed classification).

### Scoring approach for predictive modeling

The key step in predictive modeling is to construct informative (predictive) scores from the available raw data. To this end, we employed a hybrid approach that combines (a) expert-driven design of complex scores to incorporate human judgement and experience and (b) a purely data-driven approach to assigning weights to variables and then selecting the most predictive variables. The group of domain experts consisted of eight consultants with an academic life science background and several years of work experience in pharma R&D. Notably, expert input was not employed with regard to individual trials (which would lead to biased results), but exclusively in the structural design of the scoring methodology. Fig 2 provides a conceptual overview of this approach illustrated by an exemplary drug X, which is developed in three different indications A, B, and C (Fig 2A). We identified a battery of information available at a given point in time (Fig 2A, time of assessment, vertical grey line) to assess the probability of success for a given PhIII trial of interest (TOI, blue box). We classified all available information into time-dependent variables (Fig 2B, eTable 1), drug-related characteristics (Fig 2C, eTable 2) and trial-specific characteristics (Fig 2D, eTable 3). Within each of these categories we created composite scores combining weight and magnitude of information. All composites were broken down to objective and quantifiable elements (Fig 2B i-iii). We developed unbiased decision matrices for variables that offered no direct read-out from the database (e.g. Relatedness between PhII and PhIII TOI, eFig 2). In total, we created three classes of parameters comprising 16 composite scores (see eTables 2-4) based on 82 variables. Consequently, each PhIII trial is described by a unique set of descriptors, allowing the generation of a trial-specific prediction.

**Figure 2.**
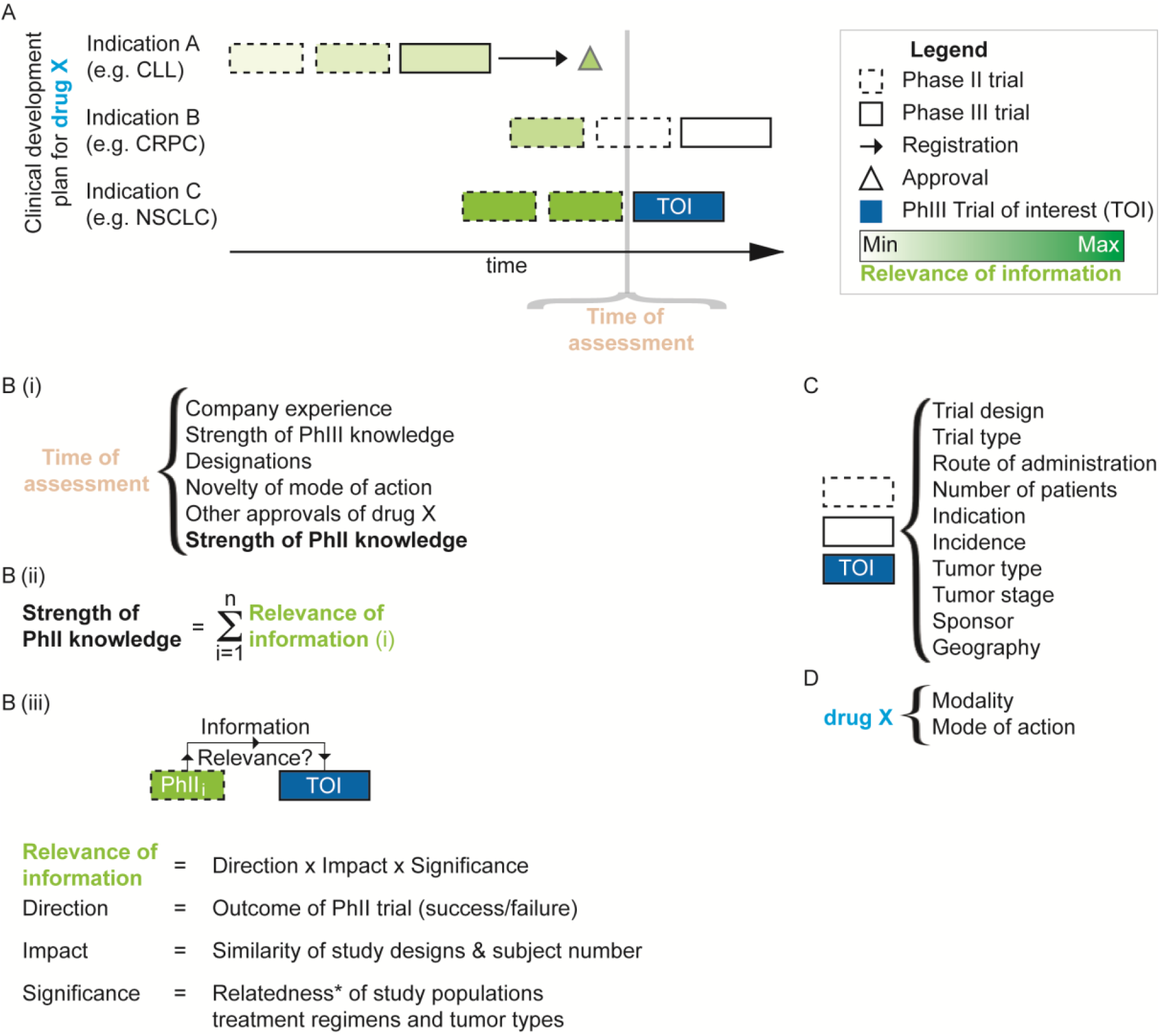
Identification, classification and weighting of variables relevant for PhIII trial assessment. (A) Schematic clinical development plan for drug X developed by sponsor Y. At time of assessment (vertical grey line) not all trials are completed (boxes with white fillings) hence cannot contribute information for assessing the PoS of the trial of interest (TOI, blue). Trials that were completed prior to the assessment carry information, but the relevance of this information (shades of green) varies with regard to the specific characteristics of the TOI. Generally speaking, the closer the patient population, the study design and the treatment algorithm of a given trial with regard to the TOI, the higher its relevance. Note, the clinical development plan (CDP) illustrates only elements, which are considered in the database. We identified three large categories of parameters: Time-dependent variables (B), trial-specific characteristics (C) and drug-related characteristics (D). (B.i) Time-dependent variables at time of assessment. Each of these variables is a composite score of several sub-variables, as illustrated by the variable Strength of PhII knowledge (SPIIK, bold). (B.ii) Weighting of relevant information exemplified by variable ‘Strength of phase II knowledge’. SPIIK is a composite of all relevant information (ROI) *i* carried by all PhII studies completed at least 2 months prior to the time of assessment. The more PhII studies *i* were completed, the stronger the body of evidence. (B.iii) Definition of ROI. Any given PhII trial *i* (green) conducted with drug X (not shown) carries information (black arrows) that are potentially relevant for assessing the PoS of the TOI (blue). The ROI for a given trial *i* is defined as the product of three factors, each of which can be broken down into further subfactors, that may even be further broken down (e.g. Relatedness, *see eFig 2 for details) until an objective and quantifiable level of information is found. Note, that there is a unique ROI for each combination of PhII and TOI, thus a unique SPIIK for each TOI. (C) Characteristics of novel therapeutic. (D) Inherent characteristics of trial of interest.

### Generation of predictive model

The overall approach to predictive modeling is a dynamic Bayesian logistic regression^23^. More specifically, the log-odds of the binary target variable (Fig 3, Outcome of TOI *S* either success or failure) is modeled as a linear function of the independent variables with a Gaussian prior for each variable *k*’s weight *α* (Fig 3A) resulting in a trial specific PoS Θ. The coefficients (weights) are estimated in a Bayesian approach using a Markov Chain Monte Carlo (MCMC)^24^ simulation to generate samples from the posterior of each parameter (Fig 3B); the final estimate is the posterior mean (Fig 3C).

**Figure 3.**
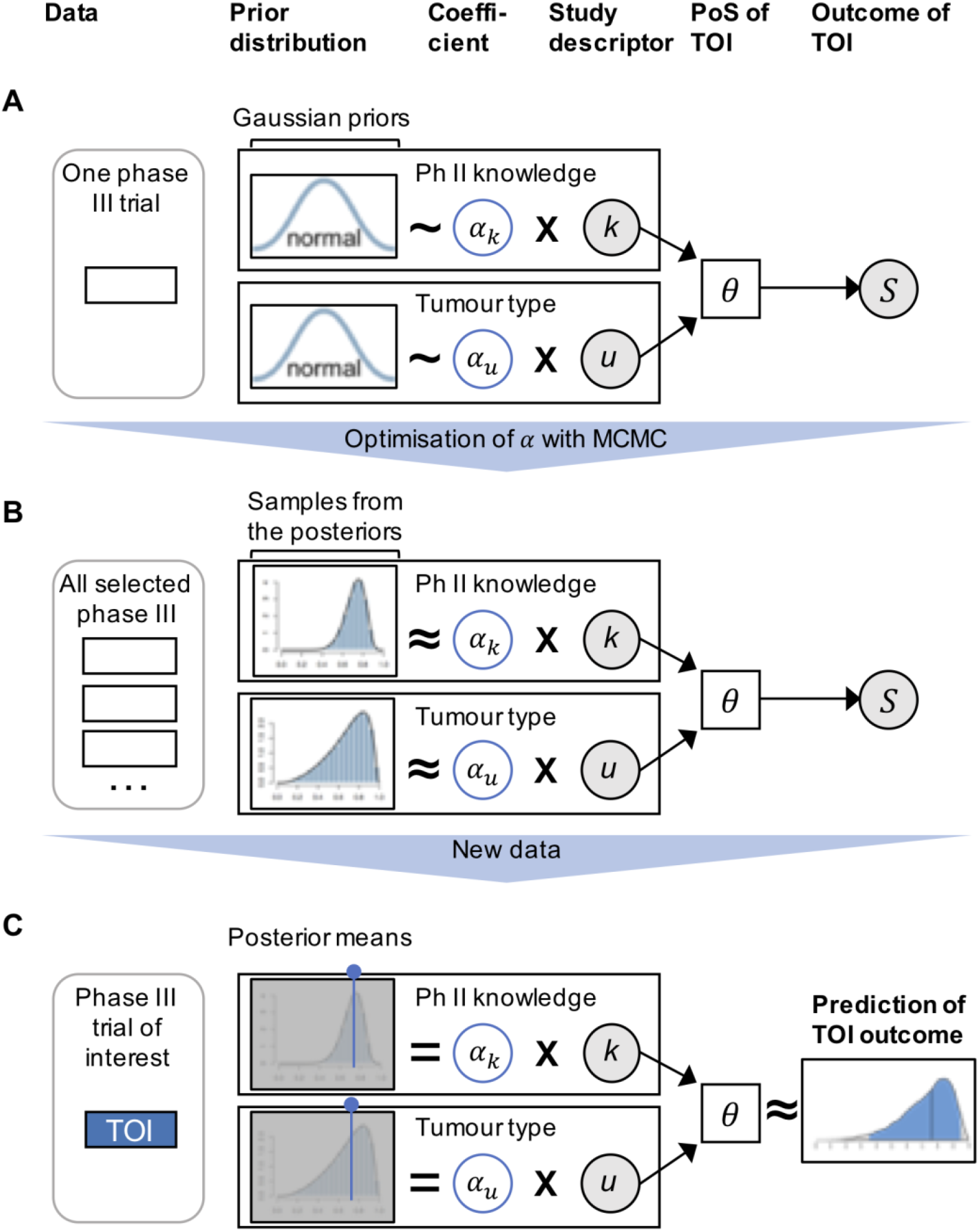
Schematic representation of Bayesian logistic regression. (A) Starting point: Bayesian model without assumptions about the parameters. (B) Model training: samples from the posteriors gained from Markov chain Monte Carlo (MCMC). (C) Calculation of PoS_TOI_ for ongoing or planned PhIII trial.

### Training and evaluation of the predictive model

As a performance metric, we use the Area under the Receiver-Operator Characteristic (AUROC)^25^. To account for the time structure in the data (PhIII trial initiation spanning 2003-2012), we use a dynamic modeling approach with regard to (a) the construction of the independent variables from raw data (Fig 4A), (b) variable selection (Fig 4B) and (c) overall performance evaluation (Fig 4C). Regarding step (a), this means that in the computation of scores only such information is used that was available at the time point of the respective PhIII trial’s initiation date (eTable 1). For variable selection (b) and overall performance evaluation (c) we adopt a time-series cross-validation strategy.

**Figure 4.**
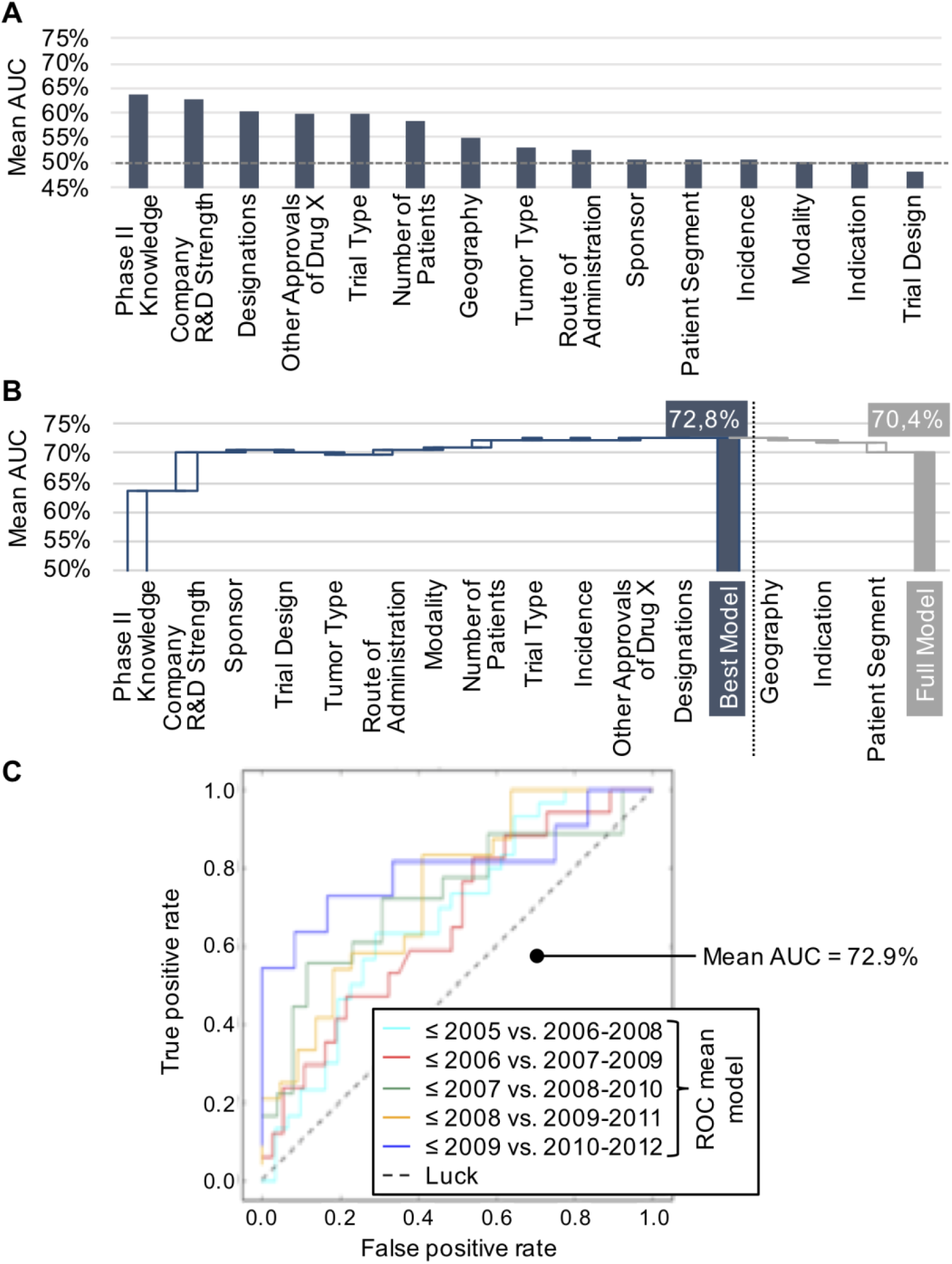
Model calibration and evaluation of performance. (A) AUC analysis of single-variables. The greater the distance from AUC = 0.5 (dotted line), the stronger the predictive performance of the variable. (B) At each step, the composite score performing best in combination with the intermediate model is selected. The predictive performance is at its maximum after the inclusion of 12 composite scores. The composite scores geography, indication, and patient segment are not included in the model because they fail to increase its performance. (C) For the best model, the receiver operating characteristic curves are displayed for each time series. The overall model performance is given by the mean AUROC over all time points and is ∼73%.

### (Ir-)Relevance of variables

The predictive performance of single composite scores can be calculated for 15 out of the 16 composite scores that we designed, i.e. all scores but novelty of MoA (Fig 4A). When applying the AUROC performance metric, we found positive correlation – mean AUROC >55%, ranging up to 64% AUROC – with success or failure for the metrics SPIIK, Company R&D strength, number of subjects in trial, designations, trial type and prior registrations of the drug in other indications.

We found no correlation – AUROC >45% and <55% -with success or failure for the metrics modality, indication, geography, and involvement of Big Pharma.

### Model selection

The best full predictive risk model (PRM) was built based on the single composite scores with a positive contribution to the model, while scores without impact were omitted (Fig 4B). To arrive at the best PRM, we added – at each step – the variable producing the highest mean AUROC increase of the model (or the smallest decrease). The variables selected (Fig 4B) are those that bring the highest overall predictive performance, reported as the mean AUROC over five time-series cross-validation splits (Fig 4C, Fig 3S). Note that the predictive performance of the single scores is not strictly additive due to overlapping information.

### Overall model performance

The performance of the model with the best combination is ∼73%_oPP_ and includes 12 variables (Fig 4C, eFig 3B, dotted line). In other words, confronted with one successful and one unsuccessful trial, using the PRM one will correctly identify the successful trial in ∼73% of the cases by picking the trial with the higher predicted PoS.

### Exemplary model outcomes

To illustrate the model’s output on single trials, we elected to highlight the variables characterizing exemplary clinical trials and to visually signify their influence on the PoS prediction (Fig 5A). Consequently, we selected two clinical studies at different ends of the spectrum, one with low mean PoS – gefitinib in treating patients with esophageal cancer that is progressing after chemotherapy, NCT01243398^26^, Fig 5B – and one with high mean PoS – trastuzumab emtansine versus capecitabine + lapatinib in participants with HER2-positive locally advanced or metastatic breast cancer (EMILIA), NCT00829166^27^, Fig 5C.

**Figure 5.**
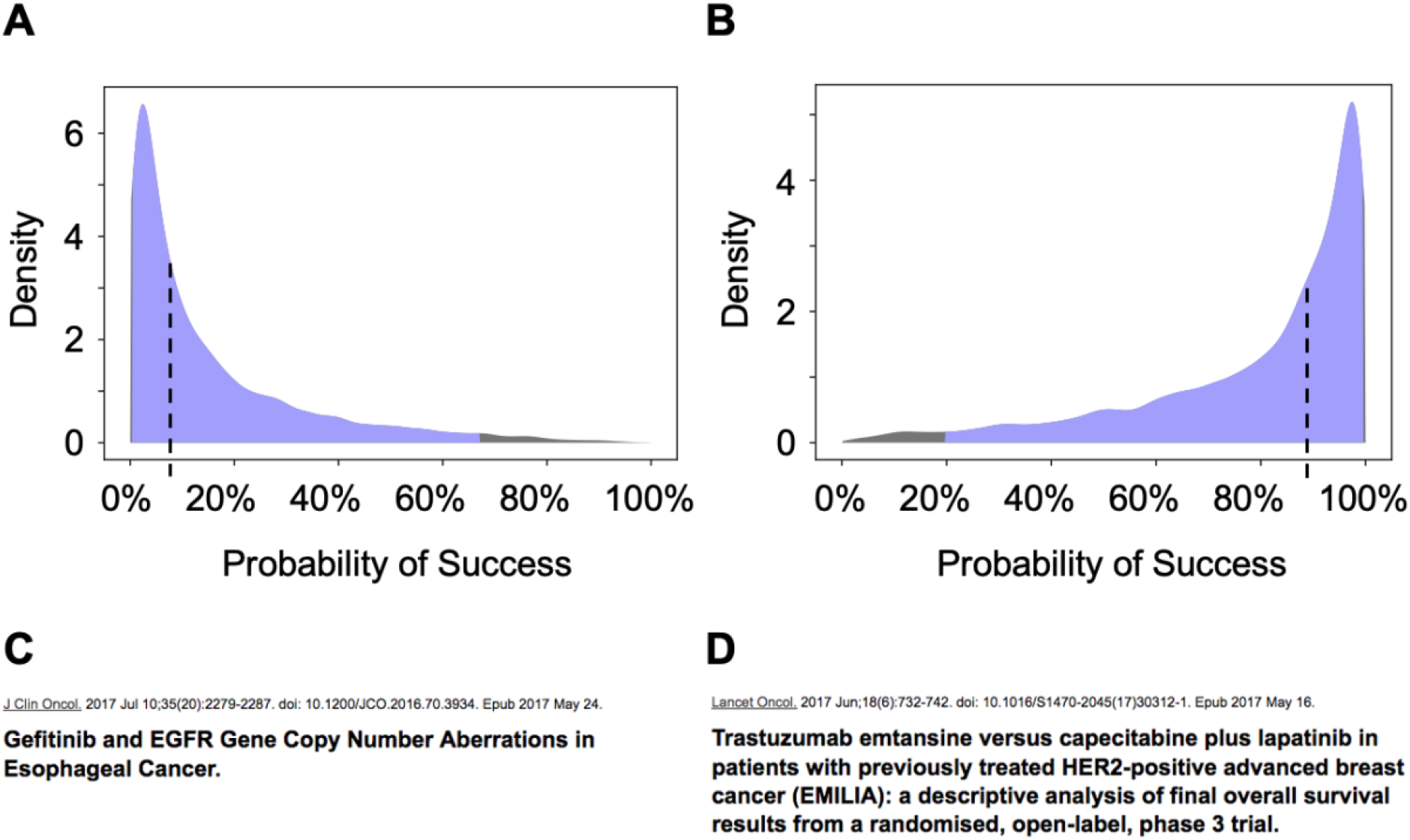
Selected PoS distributions. (A) The 13 composite scores selected for the model and the 58 variables making up those scores (unnamed for clarity) are displayed for two exemplary clinical trials. The variables characterizing each trial are marked by a dot. The variables’ weight is color coded: hue conveys the sign and intensity the amplitude. Blue indicates a positive influence on PoS prediction, red a negative one, and white no influence. (B) Study 1: The predicted PoS of the phase III trial in esophageal cancer with gefitinib (NCT01243398) is 7.8%_PoS_ (dotted line) and the 95%_CI_ credible interval ranges from 0.3%_PoS_ to 67.2%_PoS_ (colored area under the curve). (C) Study 2: The predicted PoS of the phase III trial in breast cancer with trastuzumab emtansine (NCT00829166) is 88.6%_PoS_ and the 95%_CI_ credible interval ranges from 19.6%_PoS_ to 99.6%_PoS_. The predicted PoS for those two studies are consistent with results published in peer-reviewed articles: the study with gefitinib failed (D), while the study with trastuzumab emtansine was successful (E).

The initiation dates of both studies were respectively March 2009^26^ for NCT01243398 and February 2009^27^ for NCT00829166. To mimic a real world PhIII decision point, the model goes back to the time before those particular PhIII were initiated, thereby only accessing data that were available then. The resulting PoS distributions are therefore built on 92 PhIII studies and its corresponding PhII trials preceding initiation of NCT01243398 and NCT00829166.

According to our model, the PhIII trial in in esophageal cancer with gefitinib had a predicted mean PoS_Trial_ of 7.8%_PoS_ with 95%_CI_ credible interval ranging from PoS_Trial_ = 0.3%_PoS_ to 67.2%_PoS_ at time of initiation. The study eventually failed to meet the primary endpoint median overall survival^28^ (Fig 5D). On the other hand, according to our model the PhIII trial in breast cancer with trastuzumab emtansine had a predicted mean PoS_Trial_ of 88.6%_PoS_ with 95%_CI_ credible interval ranging from PoS_Trial_ = 19.6%_PoS_ to 99.6%_PoS_ at time of initiation. That clinical study was successful^29^ (Fig 5E), confirming the predicted high probability of success.

## Discussion

Drug developers use historical success rates as forward-looking estimates to determine the PoS of an individual trial to inform their decision making. These PoS rates are often adjusted based on the opinion of subject matter experts, so-called Key Opinion Leaders (KOLs). For more than 60 years studies^30–32^ have kept demonstrating that expert opinions are no better than guessing, while in the last decade or so it was established that algorithms are able to aid or even outperform drug developers and physicians when it comes to predicting patient accrual rates in clinical trials^33,34^, optimal cancer rehabilitation, or supportive care interventions^8^. Therefore, it is surprising that an industry praising rationale drug development still takes decisions regarding investments, strategy, and science based on subjective expert advice. While the traditional approach certainly has some merit, we argue that a data-driven prediction can complement – if not quite replace – traditional methods such as KOL interviews and hSR benchmarking. In particular, we have demonstrated that the consideration of complex drivers of PhIII success such as the accumulated knowledge from prior phases is not limited to the judgement of experts (KOLs), but can also be addressed in an empirical data-driven manner using a sophisticated scoring approach presented in this study.

### Discussion of results

Employing several publicly available databases^18,21,22^ we developed a predictive model generating forward-looking and trial-specific probability of success (PoS) distributions for PhIII trials in oncology.

The most relevant single composite scores contributing to the full PRM are Strength of Ph II Knowledge (SPIIK) (mean AUROC_SPIIK_ = 64%) and Company R&D Strength at time of trial initiation (mean AUROC_SPIIK_ = 63%, Fig. 4A).

The SPIIK includes the strength and relatedness of the combined PhII evidence that exists before starting the PhIII (Fig 2, eTable 2). Our use of a decision tree optimized for prediction allowed us to model the relevance of a combined PhII body of evidence to a particular PhIII study and its design (eFig 2). The predictive performance of SPIIK alone confirms that the information building this composite score is relevant indeed. This is in line with both, common sense and regulatory guidance^35^, suggesting that the sum of PhII results are to some degree indicative for PhIII trial outcomes.

The sponsor’s past track record in oncology (Company R&D Strength) had the second largest predictive performance for PhIII studies. As this criterion includes both the number of past PhII and III studies in oncology as well as the outcomes, it essentially describes where an organization stands on the learning curve when it comes to designing studies in oncology. Noteworthy, this value is time-dependent to consider the situation on the day of trial initiation. There is a strong effect of having designed phase II and III studies that met their primary endpoints on the ability to do it again. Janssen, Celgene, and Genentech are the top 3 performers in this category of companies with at least 10 PhIII studies (2003 – 2012) in our sample.

### The role of indication

The factor ‘indication’ provides no additional value to the predictive risk model (Fig 4A, B). Notably, this is in line with expectations due to the technical bias in trial selection (Fig 1); We started our search with PhIII trials and only subsequently enriched with PhII trials associated with the selected PhIII trials. Therefore, we introduced a bias for drugs that made it into PhIII in at least one indication. Drugs exclusively developed in indications known for high failure rates (e.g. Pancreatic Cancer)^14^ hardly make it into a PhIII trial in the respective indication, hence PhIII trials in these indications are underrepresented in this proof of concept study.

### Novelty of MoA

In order to factor in the degree of innovation brought about by the compounds investigated in PhIII, we designed a composite score taking into account the novelty of MoA. That composite score was excluded from the model, as the results of our attempt were not conclusive. On one hand, this is due to the complexity of embodying the qualitative nature of innovation into a quantitative variable. On the other hand, there is a lack of availability of systematic, comprehensive data due to fundamental differences in the MoA classification schemes used and the level of information provided by companies (eTables 2 and 4).

### Comparison to other approaches

Empirical studies of clinical trial success broadly fall into two categories: (i) retrospective descriptive analysis of success rates^11,12,15,16,36,37^ and (ii) predictive approaches to modeling clinical trial success^10,38^, as in the present study. From a statistical methodology point of view, retrospective descriptive studies focus on estimates of success rates computed from empirical binary (fail vs. success)h^11,39^. Confidence intervals for the reported PoS estimates are mostly not provided, with the exception of Wong 2018^16^ (standard errors).

Among the predictive studies, a much wider range of methodological approaches can be found in the literature (eTable 5). Schachter et al.^10^ employed a Bayesian Network, a highly flexible model class, which could potentially be used to formulate expressive bottom-up generative models of trial success. Still, the approach was limited at the time due to scarcity of available databases and lack of historical data^40^, resulting in a hold-out validation set too small (n=14) to generate reliable outcomes. DiMasi *et al*. ^38^ employ an unorthodox type of predictive model which uses a scoring logic to compute the predicted PoS for a given trial.

In contrast to others, we use a linear regression model to reduce overfitting and for ease of interpretation but calibrate parameters in a Bayesian fashion so that credible intervals for parameter estimates and a posterior predictive distribution is available for PoS estimates. This is the basis for downstream Monte-Carlo simulation of portfolio-level effects. Secondly, for model evaluation we use a time-series cross-validation strategy to analyze the performance over time, as more historic information becomes available. This analysis shows not only the mean AUROC on one data set but provides also variation and stability of predictive performance.

### Limitations of approach

The current algorithm is focused on oncology PhIII trials. For this proof of concept study, we chose oncology over other therapeutic areas, because trial endpoints (mPFS and mOS) across all oncology indications are both, quantifiable and comparable in nature, providing a strong foundation for modeling approaches.

We excluded several non-standard modalities including the cellular therapies which are changing the treatment landscape as we speak. In principle, the algorithm is also prone to certain regulatory aspects (e.g. break through designations) that allow a development program to move from phase I (PhI) directly to PhIII or approval, respectively.

### Next steps & outlook

Based upon this proof of concept study, the model can potentially be expanded to (1) predict PhII trial outcomes based on data from pre-clinical and PhI studies, (2) therapeutic areas other than oncology (e.g. cardiovascular diseases), (3) incorporate more modalities (e.g. CAR T cells) for which a growing body of evidence is becoming available, and to (4) allow for the integration of non-public information available to drug developers (sponsors and investigators) in cooperation with the project teams.

## Conclusions

The algorithm presented here can distinguish successful from unsuccessful trials with much greater confidence than any other publicly available approach reviewed^10,12,14–17,38^. The positive predictive value can be tuned up to >80%_PPV_ by accepting more false negatives (lower sensitivity). To our knowledge, this is the first approach allowing to quantitatively predict the probability of success for single trials. Our model uses publicly available information only, including that of prior trials with perhaps only remote relatedness to the trial in question, and then delivers a specific prediction for a given trial. In addition, the model is fully transparent, adaptive on a trial-to-trial basis, provides unprecedented granularity (e.g. consideration of line of treatment, or background therapy) and allows identification of factors negatively (or positively) influencing the trial’s predicted PoS_Trial_.

Such an algorithm has a number of obvious applications of high medical, strategic and financial value, quite apart from the ethical dimension of a doctor’s decision to enroll patients in a study. Both sponsors and investors involved in the field of oncology could benefit greatly from a predictive algorithm assessing the prospects of a specific study, in particular by

- Supporting sponsoring companies to maximize success by designing their individual studies based on the highest possible PoS_Trial_
- Helping investors determine the impact of PhIII outcomes on valuation. This is especially relevant for those biotechs with a single PhIII asset. In addition, investors able to pursue different strategies could identify trials (and companies behind the studies) that match their investment strategy, e.g. pick-the-winner-drop-the-loser or vice versa.

## Data Availability

All data used to create this document are referenced and available at links below.

https://clinicaltrials.gov

https://www.clinicaltrialsregister.eu/ctr-search/search/

https://adisinsight.springer.com

## Acknowledgments

SH drafted manuscript and figures, developed concept, acquired, analyzed and interpreted data, managed project. MT and MK supervised project, challenged concept, provided material and technical support, and critically reviewed manuscript. LR drafted manuscript and figures, acquired, analyzed and interpreted data. JSM developed, programmed and calibrated the model, retrieved data and critically reviewed manuscript. PvB developed the model, provided statistical analysis, interpreted data, drafted the manuscript, and supervised the project.

The authors thank Birte Arlt, Nina Heid and Jennifer Price for data curation and classification, Moritz Neeb and Daniel Kirsch for advice on data architecture and for co-developing of algorithms required for model, Gerrit Buurmann for strategic advice and for expert classification of trial data, and Johannes Zimmermann for critical review of manuscript.

