## Supplementary information for "Predicting Success of Phase III Trials in Oncology"

### Supplementary Figures and Tables

### eFigure 1: Explanation of key terms used in this paper

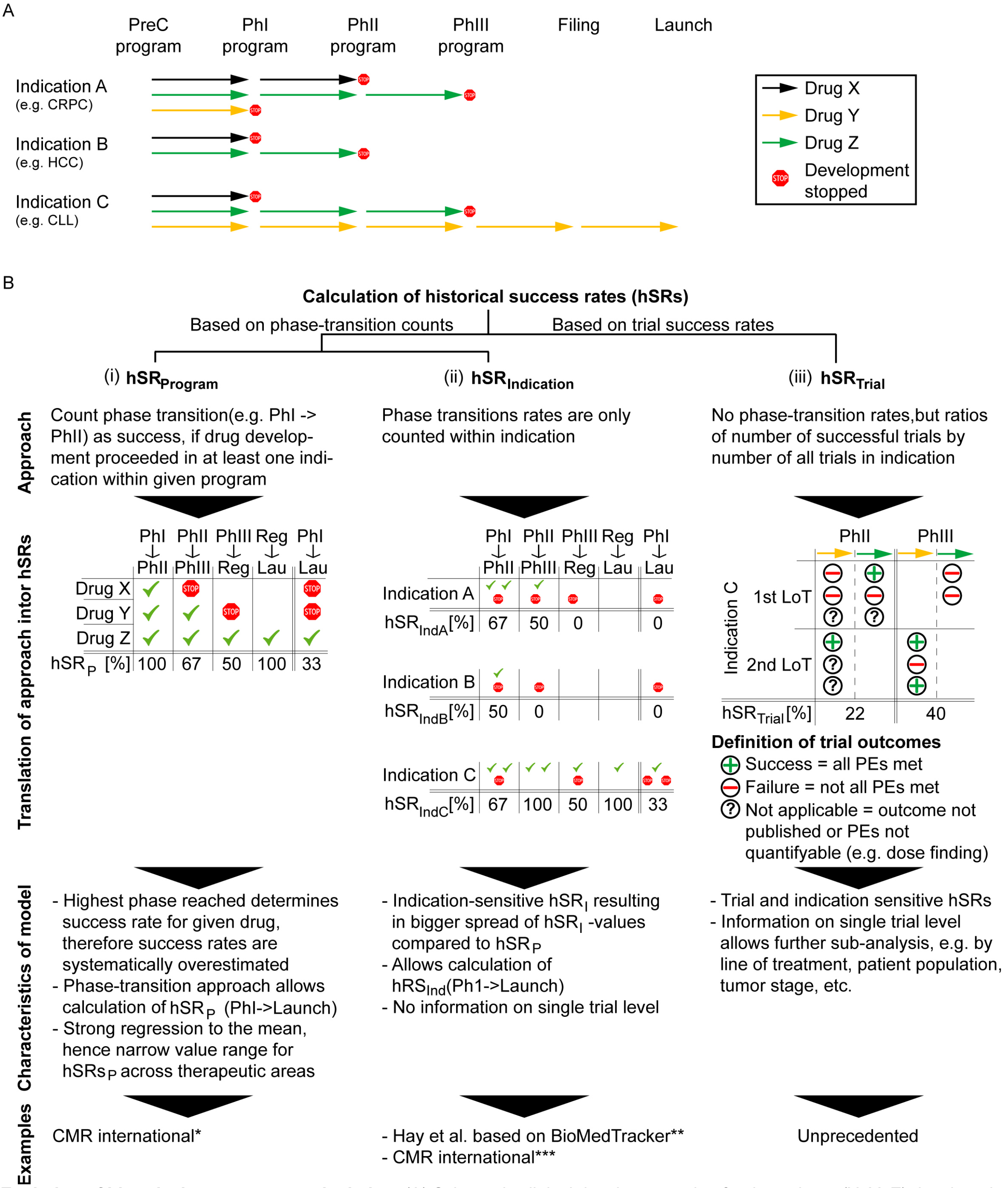


**Evolution of historical success rate calculation.** (A) Schematic clinical development plan for three drugs (X, Y, Z) developed in three indications (A, B, C). Within each indication, drug development either discontinues (stop sign) or proceeds (arrows) to the next stage of development. Only one drug (Y, yellow arrows) reaches the market (launch) in one indication (indication C). (B) Translation of schematic clinical development in (A) into historical success rates (hSRs). hSRs values for the same input data vary depending on the mode of calculation and can either be computed based on the transitions of a drug from one phase to the next ((i) and (ii)), or based on the ratio of successful trials over all trials in a given phase and indication ((iii)). Within the phase transition-based approaches, hSRs can either be calculated for programs ((i)) or for indications ((ii)). Please note, that $hSR_P$ is typically identical to the highest $hSR_I$ hence these values are somewhat comparable – though $hSR_I$s provide higher granularity than $hSR_P$s. $hSR_{Trial}$ uses a different approach and can be translated into, but not directly compared with $hSR_P$ or $hSR_I$.

*CMR pools data across related indications when companies contributing to CMR have too few projects to reach a minimal sample size per indication; **BioMedTracker uses $hSR_I$s (REF Hay et al.) as starting point/benchmark for forward-looking probability of success (PoS) calulations. The company introduces an event-based adjustment factor (EBAF [%]), which is a fixed value assigned to milestones with in a given development phase (e.g. ‘patient enrolment completed’), so that $PoS_{drug} = hSR_{I}$ + EBAF; *** CMR uses this strategy for indications with sufficient sample size (e.g. NSCLC); NSCLC = non-small cell lung cancer; HCC = hepatocellular carcinoma; CLL = chronic lymphocytic leukemia; PreC = pre-clinical drug development; hSR = historical success rate

**eFigure 2**

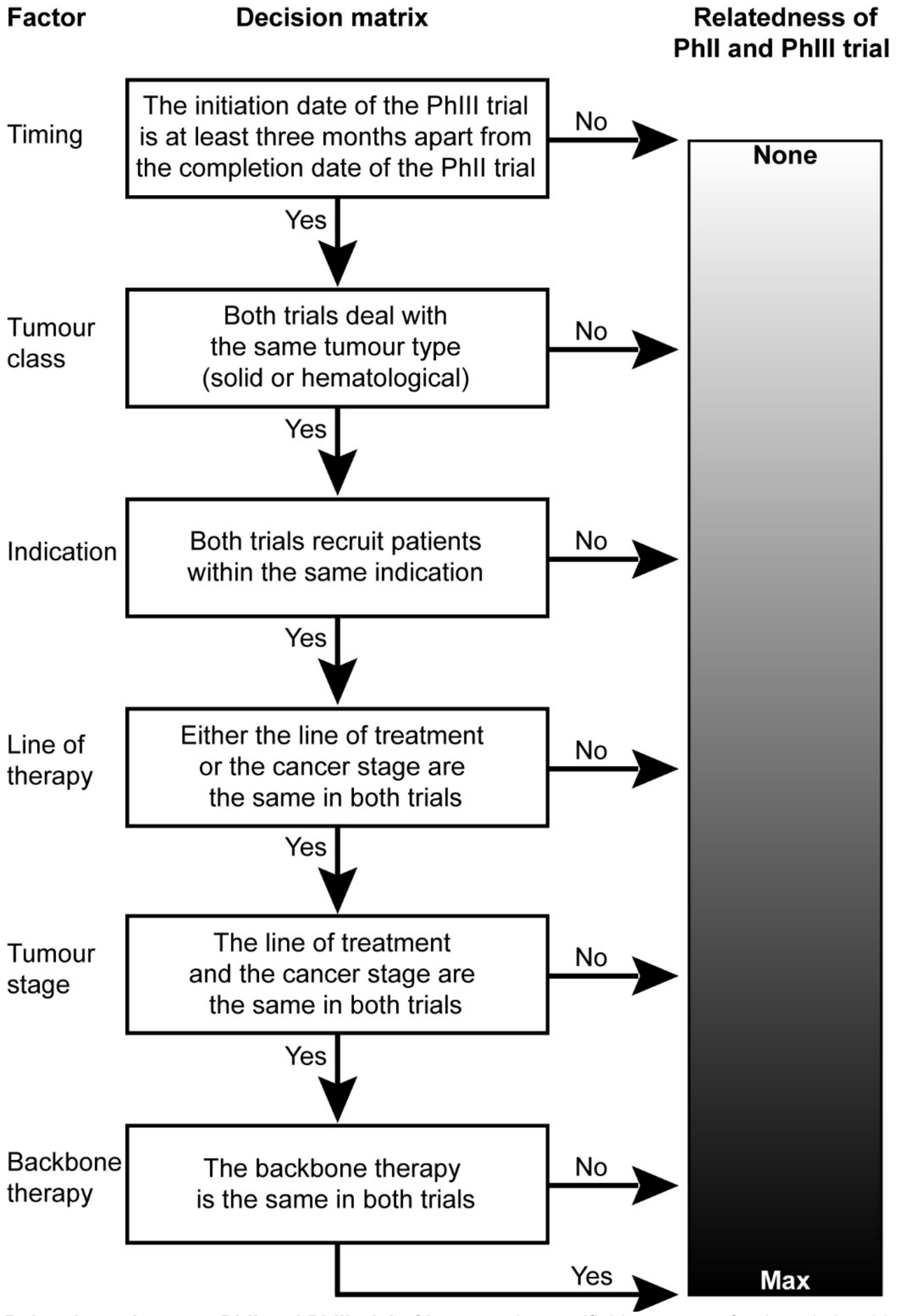


**Relatedness between PhII and PhIII trial of interest.** A quantifiable measure for the relationship was introduced by means of a decision matrix.

### eFigure 3

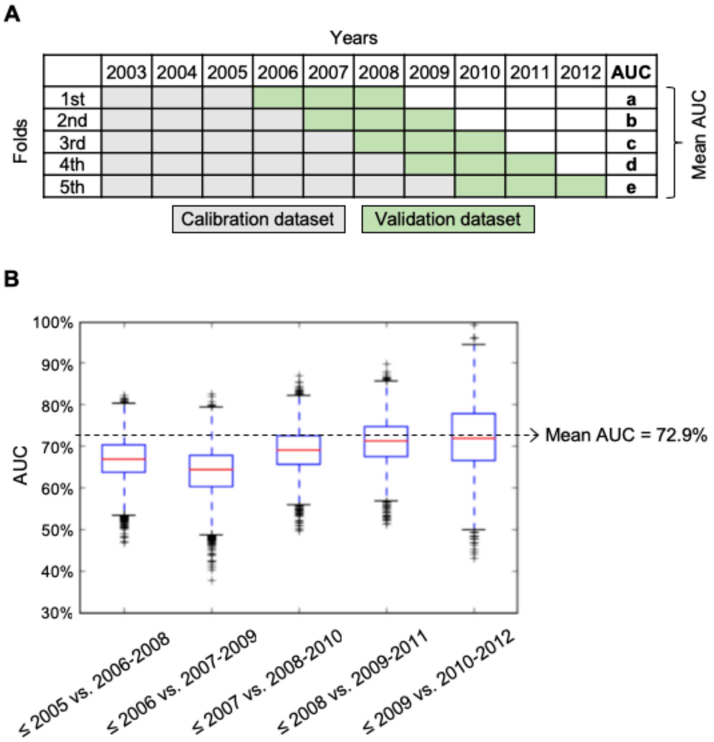


**Overall Model Performance based on Five-fold cross-validation**. (A) The overall model performance is evaluated by calculating the mean AUC on 5-fold time series cross-validation. Each fold includes three years as validation set and all previous years at training set. For this, we divide the whole data set into five temporal splits, each consisting of a pair of calibration and evaluation sets, according to the initiation dates of trials. To resemble realistic conditions in a prediction setting, the initiation dates of the trials in the calibration set precede the initiation dates of the trials in the evaluation set. Each of the five splits corresponds to a different start of the evaluation period (e.g. January 1st of 2006, 2007, 2008, 2009, and 2010 as illustrated in Fig S3A). For each split, a predictive model is estimated on the calibration set and evaluated on its corresponding evaluation set.

(B) The overall predictive performance is measured by the mean AUROC over all five splits. AUCs are calculated for each time series and sampled from the posterior distribution. Each boxplot shows the distribution of AUCs over repetitions of the time-series cross-validation at the respective point in time. The mean AUC over all time series – or overall model performance – is 72.9%.

**Tables**

### eTable 1: Explanation of key terms

| Abbreviation | Term | Unit | Definition | Comment | Applies to |
|---|---|---|---|---|---|
| **hSR** | Historical success rate | $\%_{hSR}$ | Ratio of success cases over all outcomes (see Fig S1 for details) | Typically point estimates. Often used as forward looking PoS | Depends on source and can refer to drug programs, indications, therapeutic areas, clinical trials |
| **PoS** | Probability of success | $\%_{PoS}$ | Forward-looking probability describing the rate of successful outcomes of a PhIII trial throughout all simulations and decision thresholds | Key variable of this paper: mean of the trial-specific PoS-posterior-distribution (hence confidence intervals can be provided) | Depends on sources.<br>In this paper: predicted success probability of a PhIII trial |
| **AUROC** | Area under receiver operating characteristic (often simply called AUC) | $\%_{AUROC}$ | Graphical plot illustrating the performance of a binary classifier system at varying decision thresholds (receiver-operator-curve) | Standard validation method for predictive methods for binary events. | Model |
| **PP** | Predictive performance | $\%_{PP}$ | $\%_{AUROC}$/fold | Model performance for rightly predicting the correct binary outcome across all decision thresholds in a given fold | Model |
| **oPP** | Overall predictive performance | $\%_{oPP}$ | Mean of $\%_{AUROC}$ across all time points | Average model performance for rightly predicting the correct binary outcome over all historical time points | Model |
| **DT** | Decision Threshold | $\%_{DT}$ | Threshold above which all results are classified as positive and all results below as negative | Decision point to translate PoS into binary outcome | Model |
| **PPV** | Positive predictive value | $\%_{PPV}$ | Proportion of true positive results in those that were classified positive (= above the decision threshold) | PPV is a descriptor of the model's oPP at a given DT | Model predictions |

### eTable 2: Time-dependent characteristics

| Composite Score | Description |
|---|---|
| **Company experience** | track record of the trial sponsor with regard to successfully conducted trials in the indication of interest or a related indication, respectively |
| **Strength of PhII (or PhIII) knowledge** | measured as counts of past-related PhII (or PhIII) studies, weighted by their number of subjects, study design, and endpoint overlap. Those counts qualify the PhII (or PhIII) knowledge based on the number of successes and failures |
| ***Endpoint overlap*** | Binary measure testing whether the primary endpoint – provided by the public registry – of the phase III study of interest was an endpoint in the phase II study tested. It did not matter whether it was a primary or secondary endpoint in phase II, as the information would be available for phase III regardless |
| **Designations** | fast track, orphan drug, priority review, breakthrough, conditional approval, and accelerated approval |
| **Novelty of mode of action** | score based on the number of prior studies with the MoA in question |
| **Other approvals of drug X** | registration of study drug in another patient population |

### eTable 3: Trial-specific characteristics

| Composite Score | Description |
|---|---|
| **Trial design** | quality of the design: prospective, multicenter, randomized, and double-blind |
| **Trial type** | trial masking (open label vs blinded) and purpose of trial (pivotal, exploratory, bridging, label expansion) |
| **Route of administration** | injection, oral, topical, inhalation, urogenital, rectal, other |
| **Number of patients** | number of subjects in PhIII: one of four groups optimized in width for predictive value |
| **Indication** | indications and groups of indications, e.g. colon cancer and rectal cancer studies were pooled with colorectal cancer studies |
| **Incidence** | number of newly diagnosed patients in the seven major markets (USA, Japan, UK, Germany, France, Italy, Spain) per year for the indication of interest |
| **Tumor type** | solid and hematological |
| **Patient segment** | combination of line of therapy and tumor stage; Roman numeral staging based on TNM staging system for solid tumors; no staging for hematological cancers |
| **Sponsor** | sponsor from the pharmaceutical industry, and if yes big pharma involved |
| **Geography** | location of clinical study |
| ***Classification of trial outcomes*** | Generally, only trials that meet all primary endpoints are defined as ‘successful’. Information on the trial outcomes are either retrieved from peer reviewed publications, databases, other publicly available sources or calculated from raw data where available.<br>If only raw data is provided, statistical analysis is performed based on available predetermined thresholds for significance. If trial outcomes are not publicly available, or if the outcomes lack a quantitative nature (e.g. dose-finding, RoA comparisons, ADME/PK studies) trial outcomes are classified as ‘unknown outcome’. Please note, unknown outcomes can change into either of the other classifications once new data on the trial are published.<br>Trials that do not meet all primary endpoints are counted as ‘unsuccessful’.<br>Terminated studies are also rated as ‘unsuccessful’ – unless terminated early for success – and the planned sample size is not considered |

### eTable 4: Drug-related characteristics

| Composite Score | Description |
|---|---|
| **Modality** | chemical, nucleic-acid based, protein, antibody, peptide, virus, diagnostics, cell/gene therapy, vaccine, other |
| **Mode of action** | general (e.g. immunomodulator), categorized (e.g. cytokine inhibitor) or specific (e.g. CXCL8 inhibitor) |

### eTable 5: Comparison of methodology, data and predictive performance for Schachter et al. and diMasi et al.

| | This paper | diMasi JA et al., 2015[1,2] | Schachter et al., 2007[3] |
|---|---|---|---|
| **Predicted variable** | Probability of success of individual PhIII trials in oncology | Probability of success through PhIII and marketing approval for drugs in oncology (entire programs around drugs) | Probability of success through PhIII and marketing approval for NCEs in various therapeutic areas (entire programs around drugs) |
| **Data sample** | 118 oncology drugs | 98 oncology drugs | 503 NCEs in 10 therapeutic classes, incl. 38 NCEs in oncology |
| | from the entire pharma industry | from top 50 pharma companies | from the entire pharma industry |
| | 360 PhIII studies initiated between 2003 and 2012, and 1240 PhII studies | Clinical development initiated between 1999 and 2007, and PhII recorded | Descriptive data only without information on individual NCEs |
| **Input variables** | 16 composite scores analyzed and 12 selected | >30 variables analyzed and 4 variables selected | 5 input variables feeding 2 stochastic variables |
| | Time-dependent variables (e.g. strength of PhII knowledge, sponsor's R&D experience, etc.)<br>Characteristics of drugs (i.e. modality and mode of action)<br>Characteristics of trials (e.g. number of patients, study design, etc.) | Anti-tumor activity<br>Patient number in PhII studies<br>PhII duration<br>Number of patients receiving treatment<br>intransparent | Therapeutic class<br>Development source (i.e. in-licensed vs. in-house)<br>Therapeutic indices<br>Reported data on in vitro, in vivo, and phase I/II studies<br>intransparent |
| **Model type** | Dynamic Bayesian logistic regression using MCMC simulation for posteriors | Scoring system with three categories per variable; selection of variables and tuning of category-thresholds using logistic regression followed by decision tree (CART) analysis with cross-validated AUROC as model selection criteria | Bayesian network (no information on probability distributions and parameter inference methodology provided) |
| **Validation method** | Five-fold time series cross-validation using all 360 PhIII studies | Qualitative inspection of empirical success rates in score-buckets for 61 drugs (evaluated on the calibration set) | Independent validation set of 14 NCEs in oncology |
| **Model performance (assessed by Hegge et al.)** | Mean AUROC of $73\%_{oPP}$ for the five time series | AUROC of $92\%_{AUROC}$ evaluated on the calibration set, but no separate hold-out set for validation (evaluation and calibration data sets are identical, i.e. $92\%_{AUROC}$ is not indicative of out-of-sample performance) | AUROC in the range of $61\%_{AUROC}$-$83\%_{AUROC}$ |

**Comparison of methodology, data and predictive performance for Schachter et al. and diMasi et al..** Among the prospective studies, a much wider range of methodological approaches can be found in the literature (Table S5). Schachter et al. (2007)[3] employ a Bayesian Network, a highly flexible model class, which could potentially be used to formulate expressive bottom-up generative models of trial success, directly reflecting hypothesis for the underlying causal mechanisms. However, given the scarcity of historical trial data, complex models are prone to overfitting, which can lead to (a) unreliable assessments of of predictive performance and (b) meaningless estimates of model parameters. Schachter et al. (2007)[3] do not explicitly discuss this, even though their hold-out validation set is rather small (Table S5).

Di Masi *et al.* [1] employ an unorthodox type of predictive model which uses a simple scoring logic to compute the predicted PoS for a given trial: the value of each input variable is scored independently using a maximum number of three categories; all per-variable scores are summed up to yield the final score, or PoS estimate. To calibrate this model, i.e. select variables and tune the per-variable categories and thresholds, Tufts and Janssen use a cross-validated decision tree. Due to the simple scoring logic, the model is computationally easy to apply and interpret; however, it cannot capture multivariate interactions or non-linear effects, and confidence intervals for the predictions are not available.
